# Spread of Covid-19 in the United States is controlled

**DOI:** 10.1101/2020.05.04.20091272

**Authors:** Zixin Hu, Qiyang Ge, Shudi Li, Tao Xu, Eric Boerwinkle, Li Jin, Momiao Xiong

## Abstract

As of May 1, 2020, the number of cases of Covid-19 in the US passed 1,062,446, interventions to slow down the spread of Covid-19 curtailed most social activities. Meanwhile, an economic crisis and resistance to the strict intervention measures are rising. Some researchers proposed intermittent social distancing that may drive the outbreak of Covid-19 into 2022. Questions arise about whether we should maintain or relax quarantine measures. We developed novel artificial intelligence and causal inference integrated methods for real-time prediction and control of nonlinear epidemic systems. We estimated that the peak time of the Covid-19 in the US would be April 24, 2020 and its outbreak in the US will be over by the end of July and reach 1,551,901 cases. We evaluated the impact of relaxing the current interventions for reopening economy on the spread of Covid-19. We provide tools for balancing the risks of workers and reopening economy.

## Introduction

Although as of May 1, 2020, the confirmed number cases of Covid-19 in the US has passed 1,062,446, non-pharmaceutical interventions such as strict self-quarantine for families, maintaining social distancing, stopping mass gatherings, and closure of schools and universities among others has dramatically slowed down the spread of Covid-19 and saved a large number of lives. However, public health interventions have restricted economic activities and caused high unemployment. Some investigators who published their mathematical projection of the dynamics of Covid-19 in Science suggested “prolonged or intermittent social distancing” which may drive the outbreak of Covid-19 into 2022 (1). Meanwhile, MIT researchers questioned the “intermittent social distancing” policy and worried that relaxing public interventions may cause an exponential explosion of Covid-19 (2). Now it is a critical decision point as to whether public health intervention measures should remain in place or should be lifted for reopening economy. Can we simultaneously improve both public health and economy? A key to correctly answering this question is to reconstruct the complex epidemic dynamic systems from the data, precisely predict the extent or duration of COVID-19, and develop a causal inference framework for devising practical implementable public health interventions to control the spread of Covid-19 in the US.

The basic mathematical models which underlying many statistical and computer methods for predicting the dynamics of the Covid-19 are the susceptible-exposed-infected-recovered (SEIR) models and their various versions (3-6). Although these epidemiological models are useful for estimating the dynamics of transmission, and evaluating the impact of intervention strategies, they have some critical limitations (7,8). First, the SEIR models assume a homogeneous population which is evenly mixed. Second, the epidemiological models consist of ordinary differential equations that have many unknown parameters. These parameters are not identified (9), which leads to low accuracy and a wide range of predictions. Third, most models assume that some control parameters are constant and are not time varying and system dependent. This will dramatically limit our ability to simulate interventions and improve prediction accuracy.

To overcome these limitations, we developed an artificial intelligence (AI) and causal inference integrated intervention auto-encoder (IAE) to reconstruct nonlinear time-varying epidemic dynamic systems, model health intervention plan and make multi-step predictions of the response trajectory of the Covid-19 over time with multiple interventions (fig, S1) (10). Interventions include strict travel restriction, no large group gatherings, mandatory quarantine, restricted public transportation, and school closures. Similar to reproducing number *R* in the epidemiological models, the various interventions are quantified as control variable *A_t_* taking values in the interval [0, 1]. A value of 1 for intervention indicates that intervention is the strongest and reproducing number R is close to zero. A value of zero for intervention variables indicates that no restrictions on social-economic activities are imposed. We assume that the time varying intervention variable *A_t_* is system dependent and can be automatically adjusted. As shown in Figure S1, the IAE determines the intervention response (similar to counterfactual outputs) for a set of time varying and system adjusted interventions *A_t_* and evaluates the impact of different intervention strategies and their implementation times on curbing the spread of Covid-19 and provides timely selection of an optimal sequence of intervention strategies to balance public health and economy reopening.

## Methods

### SEIR model

We first introduce the susceptible-exposed-infected-recovered (SEIR) model which is a mathematical compartmental model based on the average behavior of a population under study (1). The SEIR model is defined as

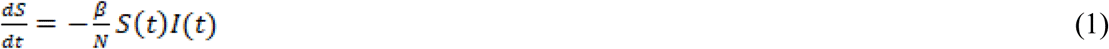

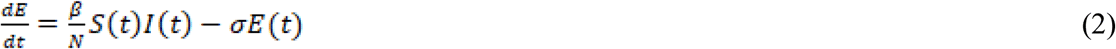

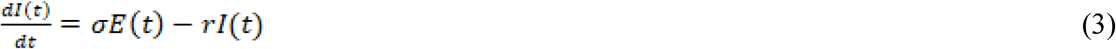

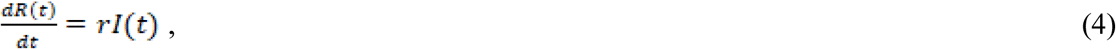

where *S*(*t*), E(t), I(t) and *R*(*t*) are the numbers of susceptible, exposed, infected and recovered (recovery or death) individuals at time *t*, respectively, *N*(*t*) is the population size, and *β*(*t*), *σ*(*t*) and *γ*(*t*) are transmission, incubation and recovery rate at time *t*, respectively.

Solving the differential equations (1)-(4), we obtain

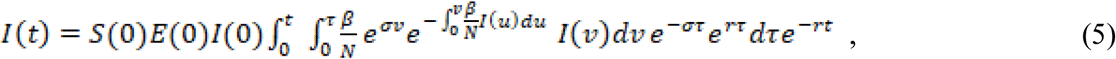

where *S*(0),*E*(0),*I*(0) and if *R*(0) are the initial values of *S*(*t*), E(t), I(t) and *R*(*t*).

For the convenience of discussion, *I*(*t*) is denoted by *Y_t_*. The observed *Y_t_* is a nonlinear function of history of *Y_t_*, parameters *β, σ* and *r*. Public health interventions such as social distancing, regional lockdowns, quarantine and intensive testing can change these parameters. In the classical SEIR and SIR model, we define the basic reproduction number as

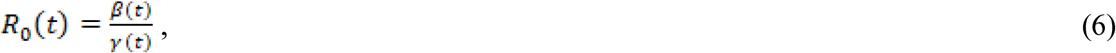

which measures the transmission dynamic properties.

The parameters *β*, *σ* and *r* depend on the time t and hence are denoted by *β*(*t*), *σ*(*t*) and *r*(*t*) Since it is difficult to quantify public health interventions, the parameters *β*(*t*), *σ*(*t*) and *r*(*t*) can also be taken as control variables. We can define a scale or vector of intervention measure *A*(*t*) to comprehensively represent the control parameters *β*(*t*), *σ*(*t*) and *r*(*t*), Equation (5) can be generally rewritten as

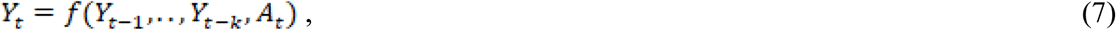

were *k* is the number of time lags.

The intervention measure *A_t_* can also be written as

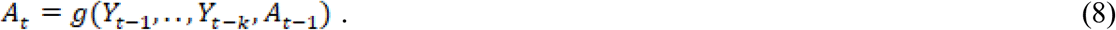

### Stacked autoencoders

Single layer autoencoder (AE) is a three layer feedforward neural network (2). The first layer is the input layer, the third layer is the reconstruction layer, and the second layer is the hidden layer. The input vector is denoted by *X_t_* = *[Y_t_, Y_t–_*_1_,*…,Y_t_*_−_*_k_*_−1,_*A_t_*]*^T^*, where *Y_t_* is the number of cases at the time *t* and 0 ≤ *A_t_ ≤* 1 is the public health intervention measure variable. The input vector is mapped to the hidden layer to capture the features of the transmission dynamics of Covid-19 with public health intervention.

AE attempts to generate an output that reconstructs its input by mapping the hidden vector to the reconstruction layer. The single layer AE attempts to minimize the error between the input vector and the reconstruction vector. We develop stacked autoencoders with 4 layers that consist of two single-layer AEs stacked layer by layer (2). The dimensions of the input layer, the first hidden layer and the second hidden layer are 8, 32 and 4, respectively (Figure S1(a)). After the first single-layer AE is trained, we remove the reconstruction layer of the first single layer AE and keep the hidden layer of the first single AE as the input layer of the second single-layer AE. Repeat the training process for the second single-layer AE. The output of the final node that fully connects to the hidden layer of the second single-layer AE is the predicted number of cases 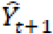 intervention measure 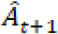.

### Potential Outcomes Framework for Evaluating the Dynamics of Covid-19 under a Sequence of Public Health Interventions

The potential outcome framework that is also referred as the Rubin Causal Model (3) is a powerful tool for modeling health intervention plan and making multi-step prediction of the response trajectory of Covid-19 over time with a sequence of public health interventions. Potential outcomes consist of observed and counterfactual outcomes. We are interested in number of cases of Covid-19 under some specific intervention. We observed the number of new cases or cumulative cases of Covid-19 (actual observation) without intervention or with some specific intervention. However, we want to know what number of new or cumulative cases of Covid-19 (counterfactual, unobserved) would be if other interventions were implemented.

Let *A_t_* be an intervention measure at time *t A_t_* − *a_t_* can be a binary variable. For example, *a_t_* = 1 (*a_t_* = 0) indicates that intervention is (not) implemented. *A_t_* = *a_t_* can also be continuous variable taking values in the interval *a_t_* ∊ [0,1]. If *A_t_* is a continuous variable, the value of *A_t_* = *a_t_* represents the intensity of intervention. *a_t_* = 1 indicates that the intervention is the most strict and comprehensive public health intervention. Let *Y_t+_*_1_ = *Y*(*a_t_*) be the potential outcome under intervention *a_t_* and be observed only when *A_t_ = a_t_* The potential outcome framework assumes the existence of the hypothetical outcome with some interventions which is not observed in the data. The hypothetical outcome under hypothetical intervention is called counterfactual outcome. The set {*A_t_*,*Y_t_*_+1_} forms a potential framework for causal inference.

### Intervention Autoencoder for Real-Time Identification, Prediction and Control of Nonlinear Time-Varying Epidemic Dynamic Systems

The IAE uses sequence-to-sequence multi-input/output architectures to model health intervention plan and make multi-step prediction of the response trajectory of Covid-19 over time with multiple interventions (Figure S1(b)). The IAE can learn the complex dynamics within the temporal ordering of input time series of Covid-19 and use an internal memory to remember. The health intervention plan has multiple intervention regimens. The IAE consists of two auto-encoders: Auto-encoder (1) is used as encoder and Auto-encoder (2) is used as the decoder. Auto-encoder (1) models input time series and a sequence of interventions (past history of the number of cases of Covid-19 and interventions over time) and predicts future response time series and interventions. Auto-encoder (2) uses the learned features of the dynamics of Covid-19 in the auto-encoder (1) to forecast the potential response time series and interventions as an input to the auto-encoder (2). The feature vector learned in the auto-encoder (1) is then provided as an input to the autoencoder (2) which initiate prediction of the future dynamics of Covid-19 under the future interventions (Figure S1(b)). The algorithm for training and forecasting of IAE is summarized as follows.

#### Algorithm

**Step 1. Initialization**.

Randomly select *A_i,t_,t* = 1,…,*T,i* = 1*,…,n* for *n* samples with *T* time points. Using the data for US and all states and regions, we train the network. Repeat above procedure five times.

**Step 2**. After the networks are trained, for each sample and each window, divide *A_i,t_* into ten grids 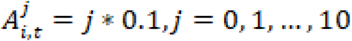. For each 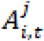, train the network:

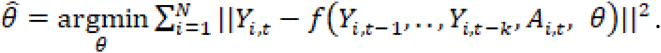

After the network is trained, for each sample, we calculate the prediction error 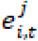.

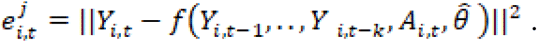

Select 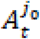 such that error is the smallest, i.e.,

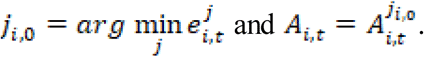

**Step 3**. Define the equation that is implemented by neural networks:

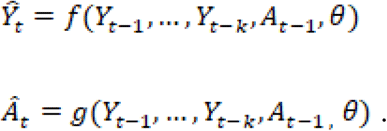

Train the network to estimate the parameters in the network, assuming that *A_t_* is estimated in step 2. In other words, we optimization the following problem:

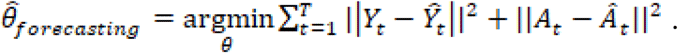

**Step 4**. Using the trained autoencoder (1) as auto-encoder (2). Predict 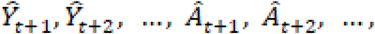 using the formula:

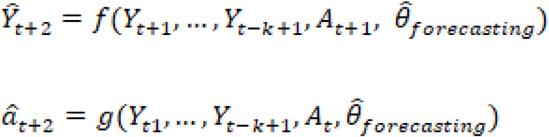

### Forecasting Procedures

The trained IAE was used to forecast the future number of new or cumulative cases of Covid-19 for US and each state. The recursive multiple-step forecasting involved using a one-step model multiple times where the prediction for the preceding time step and intervention strategy were used as an input for making a prediction on the following time step (Figure S1(b)). For example, for forecasting the number of new confirmed cases for the one more next day, the predicted number of new cases and intervention measure in one-step forecasting would be used as an observational input in order to predict day 2. Repeat the above process to obtain the two-step forecasting. The summation of the final forecasted number of new or cumulative confirmed cases for each state was taken as the prediction of the total number of new or cumulative confirmed cases of Covid-19 in US.

### Data Sources

The analysis is based on the surveillance data of confirmed and new Covid-19 cases in the US up to April 24, 2020. Data on the number of confirmed, new and death cases of Covid-19 from January 22, 2020 to April 24 were obtained from the John Hopkins Coronavirus Resource Center (https://coronavirus.jhu.edu/MAP.HTML).

### Data Pre-processing

A segment of time series with 8 days was viewed as a sample of data and *N* segments of time series was taken as the training samples. One element from the time series and intervention data matrix *Z* is randomly selected as a start day of the segment and its 7 successive days were selected as the other days to form a segment of time series. Let *n* be the index of the segment and *t_n_* be the column index of the matrix *Z* that was selected as the starting day. The *n^th^* segment time series can be represented as 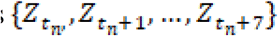. Data were normalized to 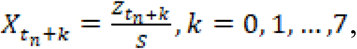 where 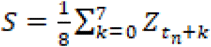. Let 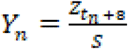 be the normalized number of new or cumulative cases to forecast. If *S* = 0, then set *Y_n_* = 0, The loss function was defined as

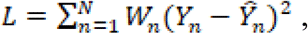

where 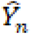 was its forecasted number of new or cumulative cases by the SAE, and *W_n_* were weights. If *t_n_* was in the interval [1, 12], then *W_n_* = 1. If *t_n_* was in the interval [13, 24], then *W_n_* = 2, etc. Repeat training processed 5 times. The average forecasting 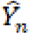 will be taken as a final forecasted number of the new or cumulative confirmed cases for each state.

## Results

### Prediction accuracy of the dynamics of Covid-19 using IAE

Accurate prediction of the spread of Covi-19 is important for future health intervention planning. To demonstrate that the IAE is an accurate forecasting method, the IAE was applied to confirmed accumulated cases of COVID-19 in the US. Fig. S2 plotted reported and one-step ahead predicted time-case curves of Covid-19 where the blue dotted curve was the number of reported cumulative cases after completion of the analysis. To further reliably evaluate the forecasting accuracy, we reported 10-step ahead forecasting errors of the cumulative cases of Covid-19 in the US, starting with April 16, 2020 (see table S1). The average errors of 1-step, 5-step and 10-step forecasting were 0.0035, 0.016 and 0.0012, respectively.

### Outbreak of Covid-19 in the US passed the peak time

We estimated that the outbreak of Covid-19 in the US would reach its peak on April 24, 2020 (Figure 1, table S2). The number of new cases and cumulative cases at peak time in the US would be 36,188 and 905,358, respectively. The forecasted number of new cases of Covid-19 in the US, 10 days after the peak would be 26,428 and drop by 27%.

**Figure 1.**
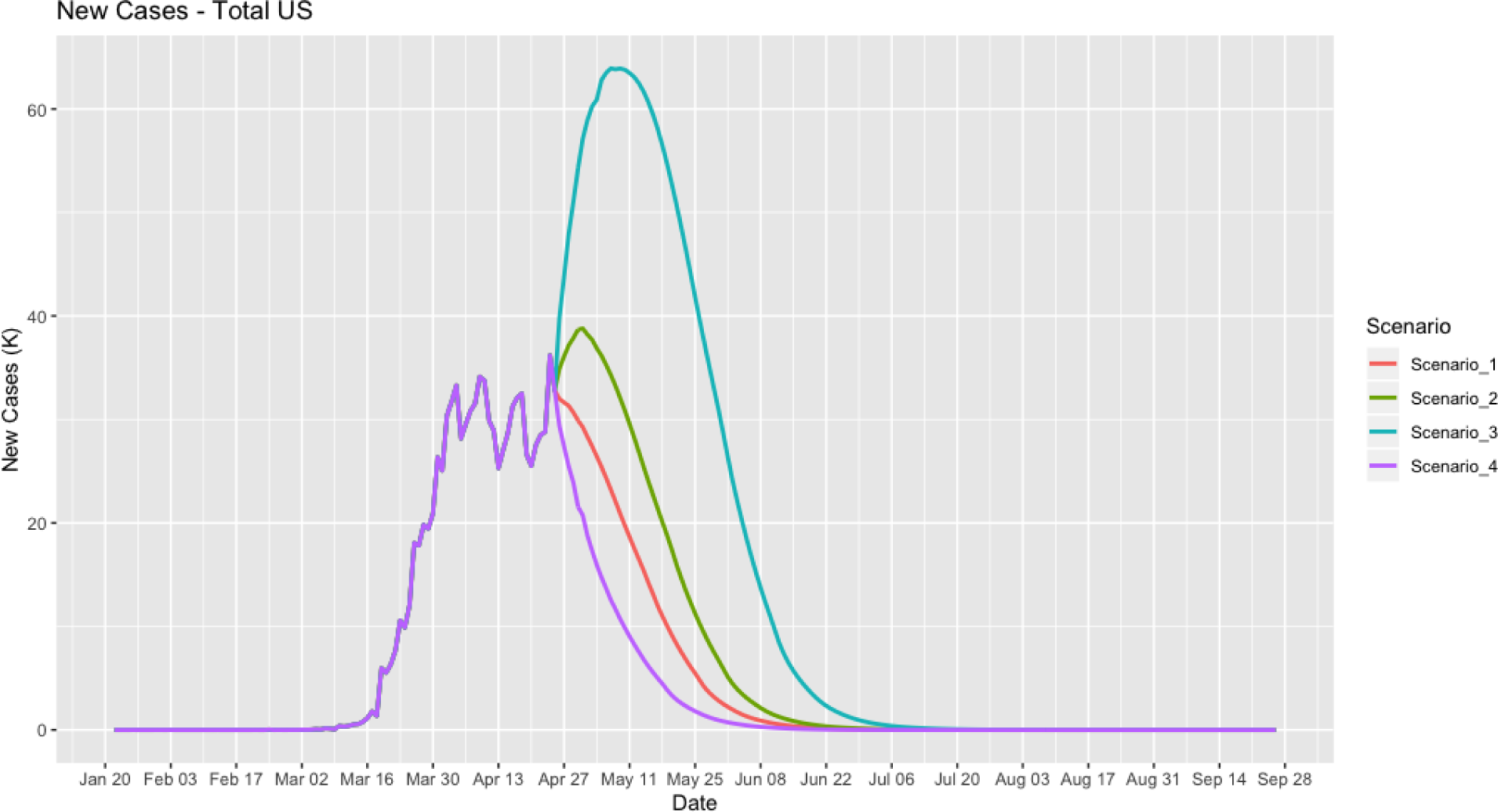
The reported and forecasted curves of newly confirmed cases of Covid-19 in the US with three scenarios of interventions as a function of time, starting date from January 22, 2020. Scenario 1 followed the current intervention measure, scenarios 2 and 3 relaxed 20% and 40% of the intervention measure, and scenario 4 increased 20% of the current intervention measure.

The peak times of the Covid-19 in the individual 50 states varied from March 24, 2020 (Virgin Islands) to May 24, 2020 (Nebraska) (Table 1 and table S2). The outbreak of the Covid-19 in New York State reached its peak on April 15, 2020 with 11,434 cases. The number of new cases and cumulative cases in the US and in the 50 individual states was summarized in Table 1. We forecasted that the outbreak of Covid-19 in the US would be completely over by the end of July. The maximum number of cases of Covid-19 in the US was 1,551,901. The end time of the outbreak of Covid-19 in the 50 states also varied from May 4 (Montana, 455 cumulative cases) to August 6, 2020 (Massachusetts, 238,370 cumulative cases). The outbreak of Covid-19 in New York State would be over on July 17, 2020 (Table 1). The maximum number of cumulative cases in New York State was 407,041. The reported and forecasted (if there are no reported) number of new cases in the US, 50 states and 5 other regions were summarized in table S2. The time - cumulative case curves of Covi-19 in the 50 states and 7 other regions were clustered into 9 groups using the k-means clustering algorithm (Figure 2). The states and regions in the same group will have the similar levels of forecasted number of cumulative cases at the end time of Covid-19.

**Figure 2.**
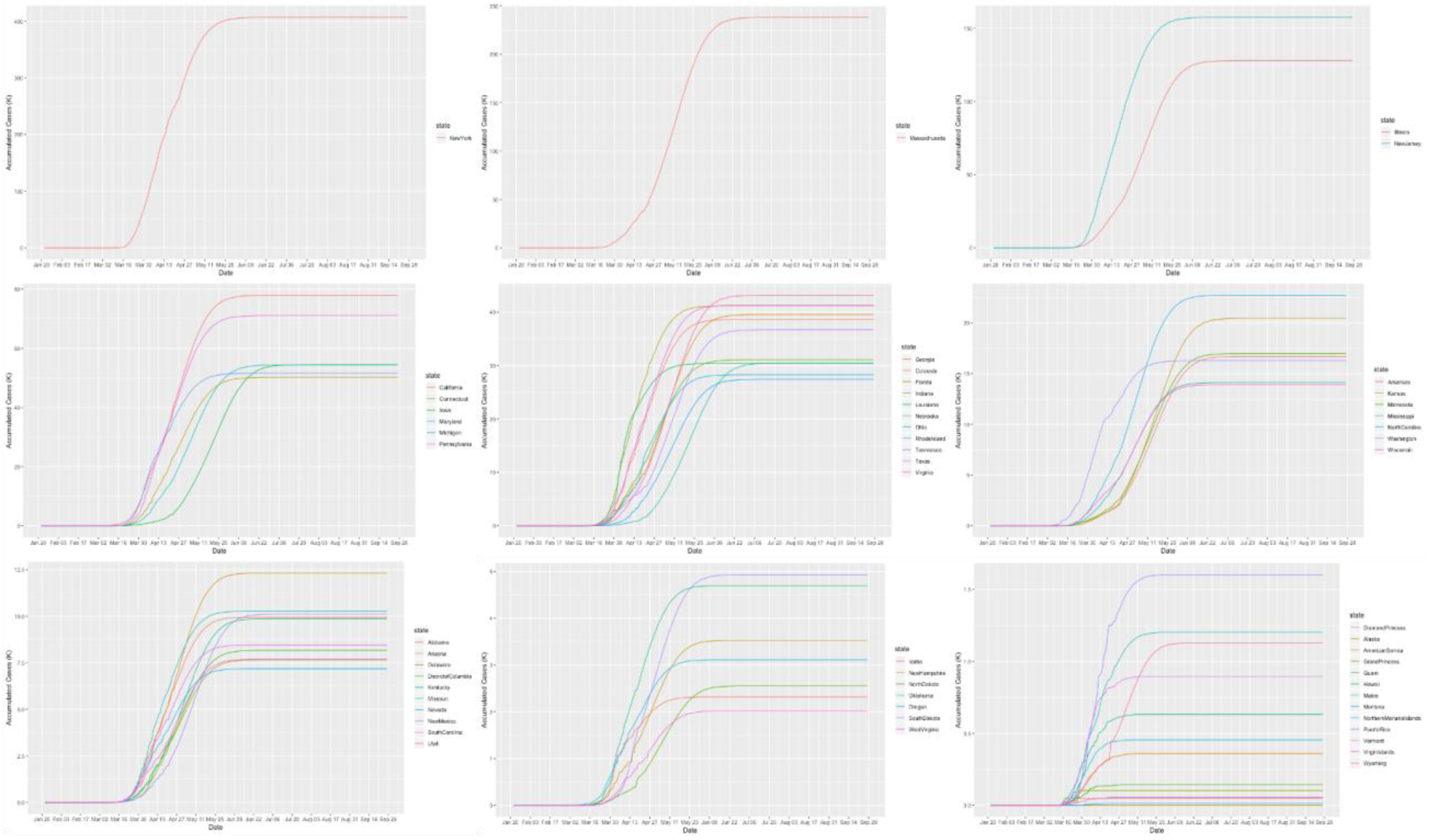
Time-case plot of 50 states. (A) Time-case plot of New York state, (B) time-case plot of Massachusetts, (C) time-case plot of New Jersey and Illinois, (D) time-case plot of California, Pennsylvania, Iowa, Maryland, Michigan and Connecticut, (E) time-case plot of Virginia, Texas, Florida, Colorado, Georgia, Tennessee, Indiana, Nebraska, Louisiana, Ohio and Rhode Island, (F) time-case plot of North Carolina, Kansas, Minnesota, Arkansas, Washington, Mississippi, and Wisconsin, (G) time-case plot of Arizona, Missouri, New Mexico, Alabama, Kentucky, South Carolina, District of Columbia, Utah, Delaware and Nevada, (H) South Dakota, Oklahoma, New Hampshire, Oregon, North Dakota, Idaho, West Virginia, and (I) time-case plot of Puerto Rico, Maine, Wyoming, Vermont, Hawaii, Montana, Alaska, Guam, Grand Princess, Virgin Islands, Diamond Princess, Northern Mariana Islands and American Samoa.

To study the impact of relaxing intervention restrictions on the spread of Covid-19 in the US, we presented the results in Figure 1. We considered four scenarios of interventions: scenario 1 followed current intervention measures, scenarios 2 and 3 relaxed 20% and 40% of the intervention measures, and scenario 4 increased 20% of the intervention measure, after April 25, 122020. Figure 1 showed that if we relaxed 40% of the intervention measure, the spread of Covid-19 would be over on August 7, with 1,869,185 cumulative cases (an increase of 317,284 cases or 20.4% of cumulative cases more than if the current intervention measure was followed) of Covid-19 in the US (table S3). To avoid increasing the number of new cases, we can increase the number of coronavirus tests.

### Intervention measures taken determines the varying time of the transmission dynamics of Covid-19

Public health interventions such as city lockdowns, traffic restrictions, quarantines, contact tracing, canceling gatherings and school closure will slow down the spread of Covid-19. Traditionally, the effects of the interventions on the transmission dynamics of Covid-19 can be investigated either by the classic SEIR epidemiological model which is determined by the exposure, infection and recovery rates *β*, *σ* and *γ*, respectively, or by the classical SIR epidemiological model which is determined by the infection rate *β* and recovery rate *γ*. The reproduction number *R_t_* in both the SEIR and SIR models is defined as

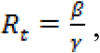

which is often used to determine the dynamic behavior of epidemics.

It is clear that information in reproduction number covers all parameters only in the SIR model and misses covering one parameter in the SEIR model. In addition, public health interventions cannot be quantified in the reproduction number. Similar to the reproducing number, we define an intervention measure *A_t_* to control the spread of Covid-19. Figure 3 plotted the intervention measure *A_t_* in the US under four scenarios of interventions as a function of times starting with January 22, 2020 and ending with the end of September, 2020. Intervention measure is a matric to quantify the degree of controlling infection. Figure 3 showed that the intervention curve started with a low intervention measure and then the trend of the intervention curve was, in general, increased until the end of February, 2020, when Spring break began. Spring break substantially reduced prevention measures and caused a large-scale outbreak of Covid-19 in the US. Then, the government implemented strict quarantine and social distance policies, and hence the intervention measure increased again. The average intervention measure of the US, 50 states and 5 other regions at the peak time was 0.54 (Table 1 and table S4). In other words, when the intervention measure was close to 0.5, interventions were sufficiently strong to decrease the number of new cases of Covid-19. Finally, the intervention measure steadily and quickly increased to 1 when the number of new cases rapidly deceased and the spread of Covid-19 was completely stopped. Figure 3 also showed that even if the intervention measure was assumed to decrease 40%, the intervention measure could still quickly and steadily increase to 1 and then the spread of Covid-19 would stop.

**Figure 3.**
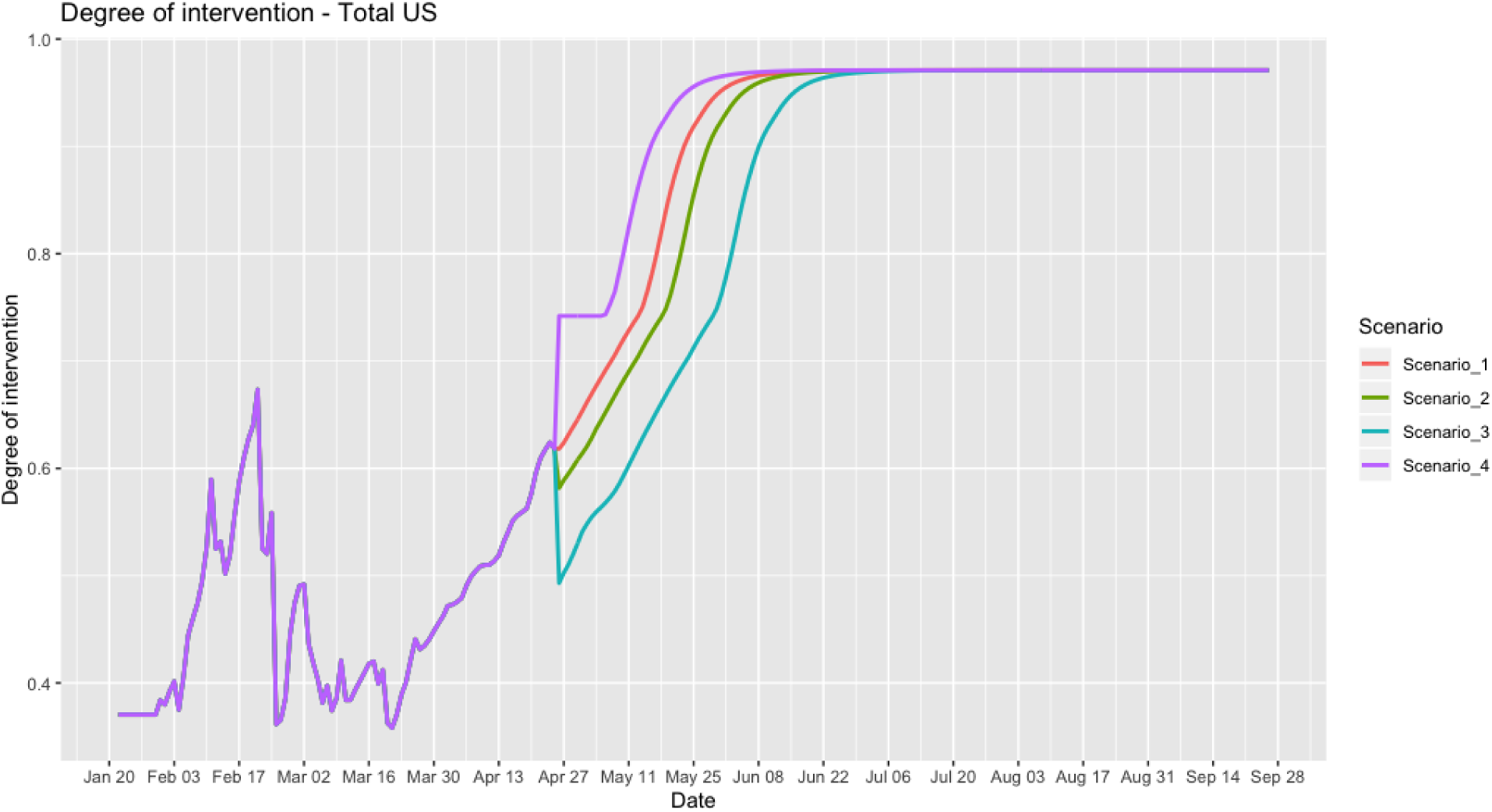
The intervention measure curves as a function of times under three scenarios of interventions to control the transmission dynamics of Covid-19 in the US.

Table S5 presented correlation coefficients between the number of new cases and the intervention measure in the US, 50 states and 5 other regions. The correlation coefficients between the number of new cases and the intervention measure in the US is −0.4819. A total of 23.2% of the variation of new cases were explained by the intervention measure. Correlation coefficients of the 50 states ranged from −0.4473 (California) to −0.1480 (Northern Mariana Islands). California (−0.4473), Washington (−0.4197), Arizona (−0.4008), Illinois (−0.3759) and Massachusetts (−0.3521) were the top five states with the largest correlation coefficients. Negative correlation coefficients indicated that increasing the intervention measure would decrease the number of new cases. To investigate the relationship between the intervention measure and widely used reproduction number R(t), we first used SIR model to calculate the reproduction number R(t) and then calculate the Spearman correlation coefficient between the intervention measure and reproduction number R(t). We obtain the Spearman correlation coefficient of 0.585 between the intervention measure and reproduction number, using the number of new cases in the US and 50 states from April 1 to April 29.

## Discussion

In summary, this report have addressed several important issues in forecasting the transmission dynamics of Covid-19 and evaluating the effects of the intervention measures on the curbing spread of Covid-19. First issue is low forecasting accuracy of the classical epidemiological models due to the unidentifiability of the model parameters. The classical epidemiological models often give a wide range of predictions of the future trajectories of the epidemics, which causes difficulties for public health intervention planning. As an alternative to epidemiologic mode, we developed data driven IAE for real-time prediction and control of nonlinear time-varying epidemic dynamic systems. The prediction accuracy of the IAE was very high. The 10-step forecasting error of IAE was 0.0012.

Second issue is how to formulate the real-time forecasting and designing intervention strategies for controlling the spread of Covid-19 as a causal inference problem. The data collected for Covid-19 are observational data. It is infeasible to collect the transmission dynamics data from the experiments. These data for both the total US and individual state can be observed only once. Dynamic responses of epidemics under multiple intervention scenarios are counterfactual. Unlike model-based approach where the models are assumed underlying the transmission dynamics of epidemics, the data driven evaluations of intervention strategies require causal inference as a basic tool for forecasting and evaluating the dynamics of Covid-19 in the US. We used counterfactual outcome as a general framework for modeling health intervention plan and making multi-step prediction of the response trajectory of Covid-19 over time with a sequence of public health interventions. As illustration, we evaluated four scenarios of interventions and predicted that if we relaxed 40% of intervention measure, the spread of Covid-19 would be over on August 7, with 1,869,185 cumulative cases (increased 317,284 cases or 20.4% of cumulative cases than following the current intervention measure) of Covid-19 in US. However, if we increased 20% of the intervention measure, for example, by increasing the ratio of coronavirus tests, the spread of Covid-19 would be over on July 23, with 1, 296,487 cumulative cases (reduced 16.5% of cumulative cases than following the current intervention measure).

The third issue is to simultaneously estimate the trajectory of the dynamics of Covid-19 and the intervention measure. We proposed to use intervention measure as a control variable that comprehensively quantified the public health interventions and incorporate the intervention measure as an input into the IAE model. Therefore, the IAE model jointly estimate the number of cases and intervention measure.

The four issue is interpretation of intervention measure. We could not investigate the impact of all individual elements of the interventions because many were introduced simultaneously across the US. If the individual intervention data are available, the IAE model can quantify the effect of the specific intervention on the controlling spread of Covid-19. The widely used quantity to characterize the transmission of dynamics is the reproduction number R. We found that the correlation coefficient between the intervention measure and reproduction number was 0.585.

The US passed the peak time of the Covid-19 and the number of new cases decreasing. The interventions such as stay-at-home orders and business closures dramatically slowed down the spread of Covid-19, but at the cost of economy shutdowns. A key to safely reopening the economy is massive virus tests. Relaxing quarantine, self-isolation and business closure is offset by increasing the number of tests. Question is how many number of tests is needed to ensure the curbing the spread of Covid-19 without intriguing the second wave of the outbreak. The IAE model with the ratio of the test as input provides tools for evaluating a sequence of strategies for safely reopening the economy.

## Data Availability

All data is published by CSSE at Johns Hopkins University at https://github.com/CSSEGISandData/COVID-19/tree/master/csse_covid_19_data

## Funding

Dr. Li Jin was partially supported by National Natural Science Foundation of China (91846302).

## Supplementary Figure

**Figure S1.**
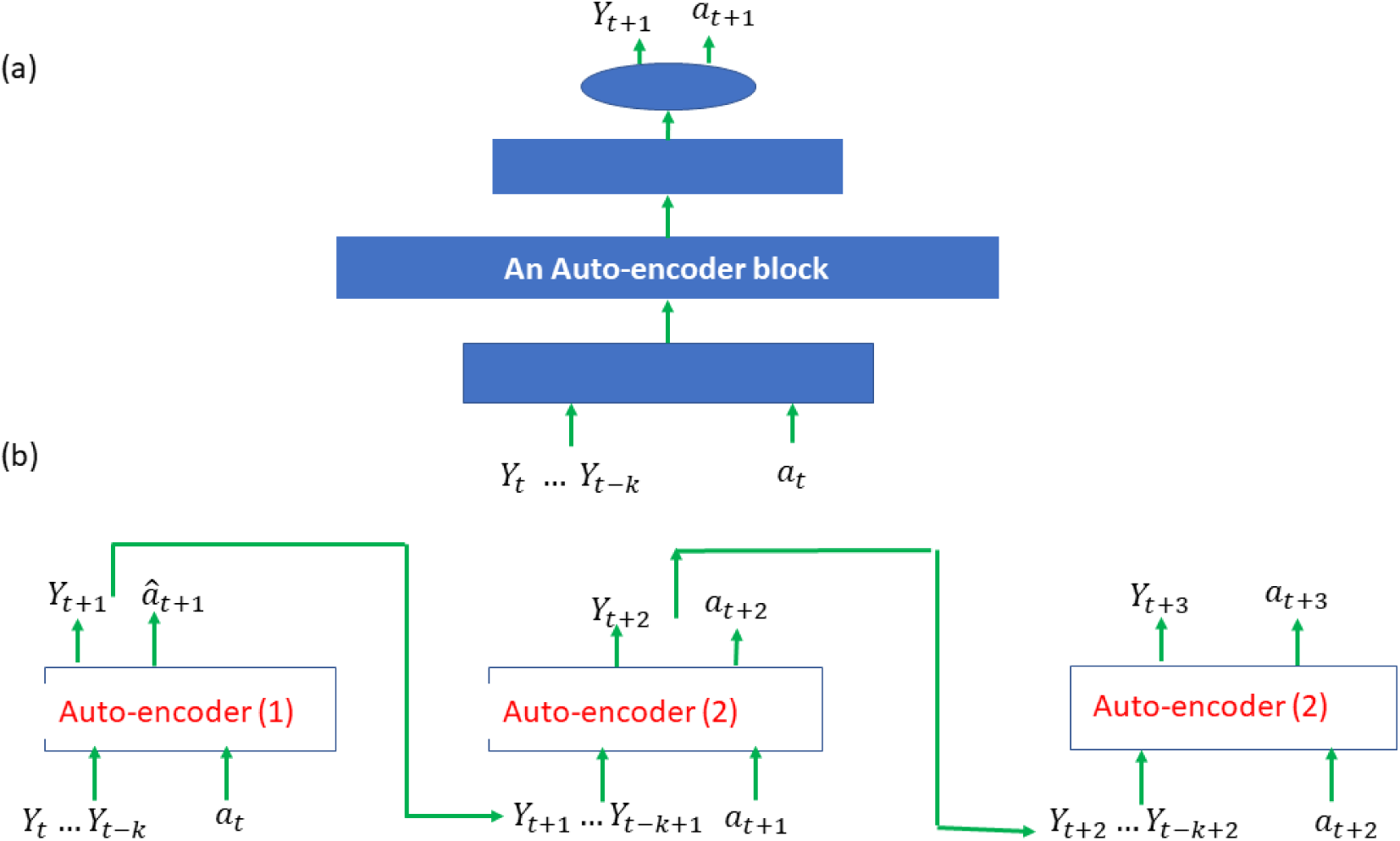
Architecture of intervention autoencoder, (a) block autoencoder with 8×32×4×2 dimensions and (b) Process structure of the developed intervention autoencoder model.

**Figure S2.**
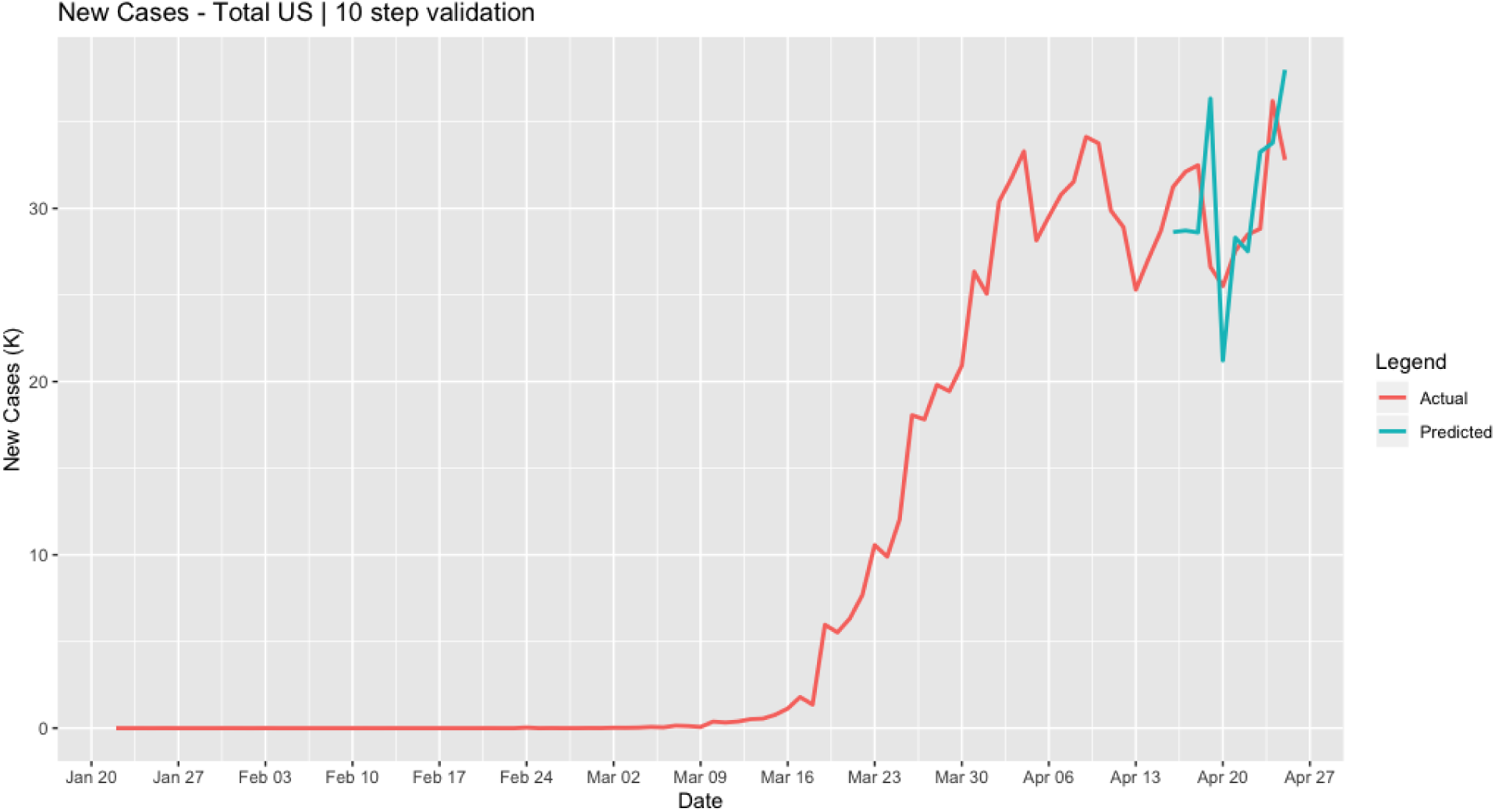
Plotted reported and one-step ahead predicted time-case curves of Covid-19 in the US.

---

**Table S1.**
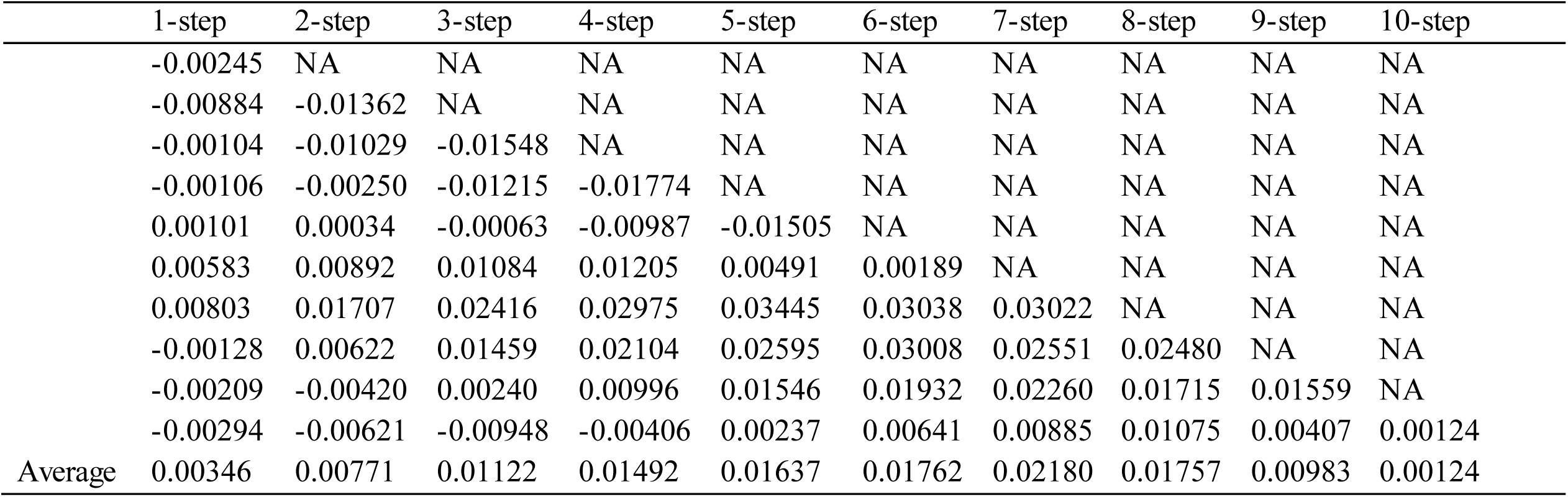
Errors of the 10-Step ahead predicting the number of cumulative cases of Covid-19 in the US.

**Table S2.**
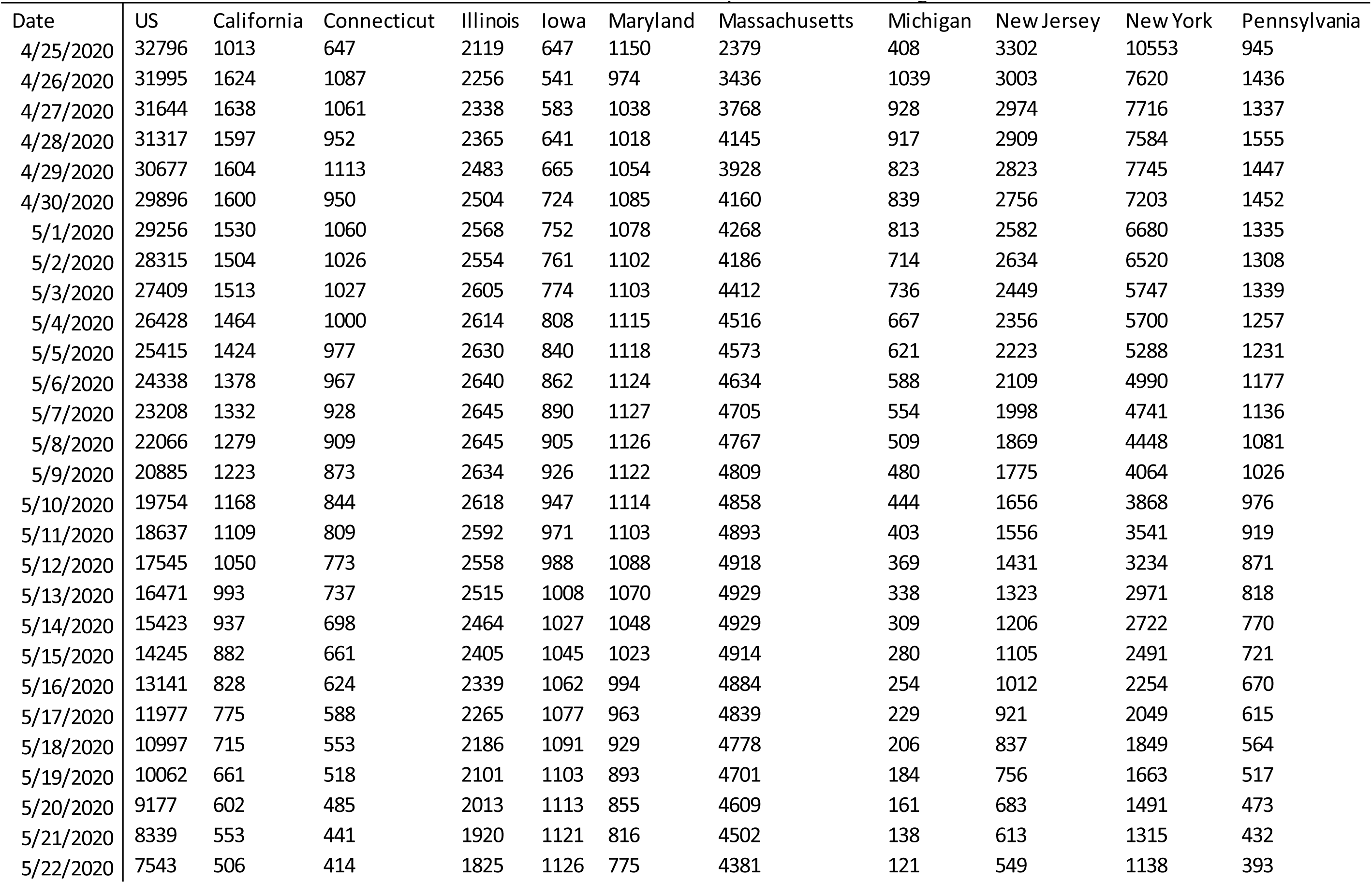
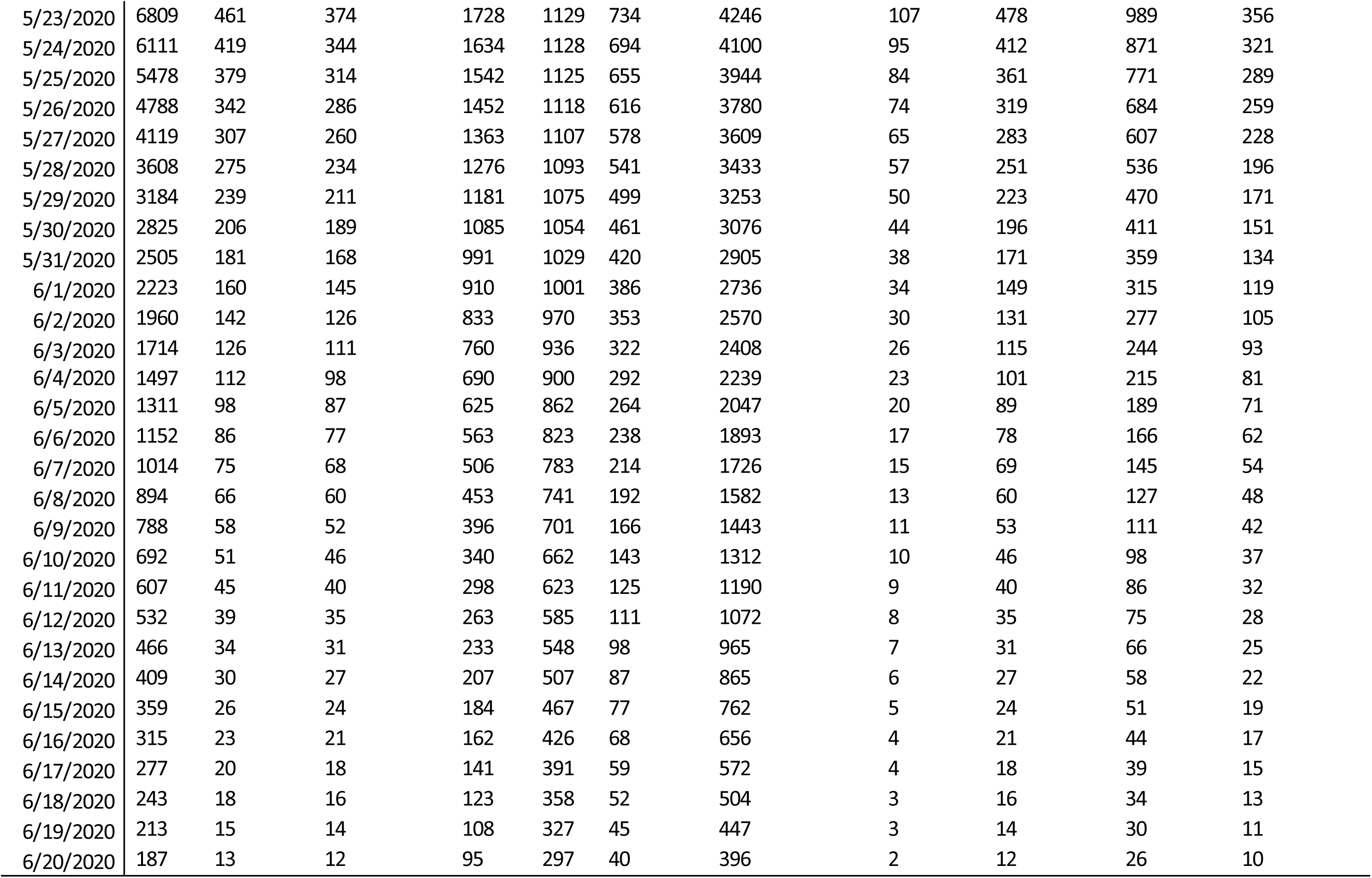
Forecasted number of new cases for 8 weeks in the US and Top States with the largest number of cumulative cases.

**Table S3.** Peak time and peak number of cases, end time and end number of cases in the US and 50 states and 7 other regions.

| Total US and State | Current Number of Cases | scenario | Peak Time | Peak Number (New) | Peak Number (Cum) | End Time | End Number of Cases |
| --- | --- | --- | --- | --- | --- | --- | --- |
| US | 938154 | Scenario 1 | 4/24/2020 | 36188 | 905358 | 7/31/2020 | 1551901 |
|  |  | Scenario 2 | 5/1/2020 | 38792 | 1161618 | 8/7/2020 | 1869185 |
|  |  | Scenario 3 | 5/7/2020 | 63920 | 1602278 | 8/22/2020 | 3083138 |
|  |  | Scenario 4 | 4/24/2020 | 36188 | 905358 | 7/23/2020 | 1296487 |
| Alabama | 6026 | Scenario 1 | 4/9/2020 | 375 | 2703 | 6/23/2020 | 9949 |
|  |  | Scenario 2 | 4/9/2020 | 375 | 2703 | 6/29/2020 | 11693 |
|  |  | Scenario 3 | 4/9/2020 | 375 | 2703 | 7/10/2020 | 17190 |
|  |  | Scenario 4 | 4/9/2020 | 375 | 2703 | 6/16/2020 | 8507 |
| Alaska | 339 | Scenario 1 | 3/28/2020 | 27 | 85 | 5/9/2020 | 360 |
|  |  | Scenario 2 | 3/28/2020 | 27 | 85 | 5/9/2020 | 360 |
|  |  | Scenario 3 | 3/28/2020 | 27 | 85 | 5/13/2020 | 375 |
|  |  | Scenario 4 | 3/28/2020 | 27 | 85 | 5/2/2020 | 347 |
| Arizona | 6286 | Scenario 1 | 4/23/2020 | 299 | 5772 | 6/29/2020 | 12316 |
|  |  | Scenario 2 | 5/3/2020 | 333 | 8766 | 7/7/2020 | 15687 |
|  |  | Scenario 3 | 5/15/2020 | 617 | 17026 | 7/23/2020 | 29767 |
|  |  | Scenario 4 | 4/23/2020 | 299 | 5772 | 6/21/2020 | 9927 |
| Arkansas | 2911 | Scenario 1 | 5/16/2020 | 347 | 9445.5 | 7/20/2020 | 16705 |
|  |  | Scenario 2 | 5/21/2020 | 522 | 14270 | 7/28/2020 | 25175 |
|  |  | Scenario 3 | 5/27/2020 | 937 | 25765.5 | 8/7/2020 | 45227 |
|  |  | Scenario 4 | 4/23/2020 | 323 | 2599 | 7/6/2020 | 9426 |
| California | 42368 | Scenario 1 | 4/20/2020 | 2255 | 33686 | 7/11/2020 | 77978 |
|  |  | Scenario 2 | 4/20/2020 | 2255 | 33686 | 7/20/2020 | 101651 |
|  |  | Scenario 3 | 5/16/2020 | 4153 | 116029 | 8/7/2020 | 200917 |
|  |  | Scenario 4 | 4/20/2020 | 2255 | 33686 | 7/5/2020 | 66237 |
| Colorado | 12968 | Scenario 1 | 4/24/2020 | 978 | 12256 | 7/17/2020 | 39566 |
|  |  | Scenario 2 | 5/14/2020 | 1260 | 34000 | 7/28/2020 | 60834 |
|  |  | Scenario 3 | 5/23/2020 | 2414 | 66319 | 8/10/2020 | 116663 |
|  |  | Scenario 4 | 4/24/2020 | 978 | 12256 | 7/4/2020 | 25995 |
| Connecticut | 24583 | Scenario 1 | 4/22/2020 | 2109 | 22469 | 7/11/2020 | 50220 |
|  |  | Scenario 2 | 4/22/2020 | 2109 | 22469 | 7/18/2020 | 64210 |
|  |  | Scenario 3 | 5/17/2020 | 2779 | 76206 | 8/6/2020 | 134409 |
|  |  | Scenario 4 | 4/22/2020 | 2109 | 22469 | 7/3/2020 | 41259 |
| Delaware | 3576 | Scenario 1 | 4/22/2020 | 269 | 3200 | 6/28/2020 | 7659 |
|  |  | Scenario 2 | 4/22/2020 | 269 | 3200 | 7/7/2020 | 10402 |
|  |  | Scenario 3 | 5/19/2020 | 462 | 12787 | 7/25/2020 | 22264 |
|  |  | Scenario 4 | 4/22/2020 | 269 | 3200 | 6/19/2020 | 6075 |
| Diamond Princess | 49 | Scenario 1 | 3/17/2020 | 47 | 47 | 3/20/2020 | 49 |
|  |  | Scenario 2 | 3/17/2020 | 47 | 47 | 3/20/2020 | 49 |
|  |  | Scenario 3 | 3/17/2020 | 47 | 47 | 3/20/2020 | 49 |
|  |  | Scenario 4 | 3/17/2020 | 47 | 47 | 3/20/2020 | 49 |
| District of Columbia | 3699 | Scenario 1 | 4/8/2020 | 229 | 1440 | 6/29/2020 | 8166 |
|  |  | Scenario 2 | 5/7/2020 | 240 | 6482 | 7/9/2020 | 11520 |
|  |  | Scenario 3 | 5/19/2020 | 492 | 13496 | 7/26/2020 | 23712 |
|  |  | Scenario 4 | 4/8/2020 | 229 | 1440 | 6/19/2020 | 6260 |
| Florida | 30839 | Scenario 1 | 4/2/2020 | 2052 | 9008 | 6/26/2020 | 41224 |
|  |  | Scenario 2 | 4/2/2020 | 2052 | 9008 | 7/1/2020 | 45202 |
|  |  | Scenario 3 | 4/2/2020 | 2052 | 9008 | 7/9/2020 | 55868 |
|  |  | Scenario 4 | 4/2/2020 | 2052 | 9008 | 6/21/2020 | 38022 |
| Georgia | 23222 | Scenario 1 | 4/17/2020 | 1525 | 17194 | 7/3/2020 | 38652 |
|  |  | Scenario 2 | 4/17/2020 | 1525 | 17194 | 7/10/2020 | 47268 |
|  |  | Scenario 3 | 5/8/2020 | 1677 | 41702 | 7/26/2020 | 80409 |
|  |  | Scenario 4 | 4/17/2020 | 1525 | 17194 | 6/25/2020 | 32650 |
| Grand Princess | 103 | Scenario 1 | 3/28/2020 | 75 | 103 | 3/28/2020 | 103 |
|  |  | Scenario 2 | 3/28/2020 | 75 | 103 | 3/28/2020 | 103 |
|  |  | Scenario 3 | 3/28/2020 | 75 | 103 | 3/28/2020 | 103 |
|  |  | Scenario 4 | 3/28/2020 | 75 | 103 | 3/28/2020 | 103 |
| Guam | 141 | Scenario 1 | 4/5/2020 | 19 | 112 | 4/28/2020 | 144 |
|  |  | Scenario 2 | 4/5/2020 | 19 | 112 | 4/28/2020 | 144 |
|  |  | Scenario 3 | 4/5/2020 | 19 | 112 | 4/28/2020 | 144 |
|  |  | Scenario 4 | 4/5/2020 | 19 | 112 | 4/27/2020 | 142 |
| Hawaii | 605 | Scenario 1 | 4/3/2020 | 63 | 319 | 5/10/2020 | 631 |
|  |  | Scenario 2 | 4/3/2020 | 63 | 319 | 5/12/2020 | 642 |
|  |  | Scenario 3 | 4/3/2020 | 63 | 319 | 5/16/2020 | 663 |
|  |  | Scenario 4 | 4/3/2020 | 63 | 319 | 5/8/2020 | 626 |
| Idaho | 1887 | Scenario 1 | 4/2/2020 | 210 | 776 | 6/1/2020 | 2318 |
|  |  | Scenario 2 | 4/2/2020 | 210 | 776 | 6/4/2020 | 2474 |
|  |  | Scenario 3 | 4/2/2020 | 210 | 776 | 6/11/2020 | 2822 |
|  |  | Scenario 4 | 4/2/2020 | 210 | 776 | 5/26/2020 | 2161 |
| Illinois | 41777 | Scenario 1 | 4/24/2020 | 2721 | 39658 | 7/26/2020 | 127855 |
|  |  | Scenario 2 | 5/13/2020 | 3822 | 104683 | 8/4/2020 | 184875 |
|  |  | Scenario 3 | 5/23/2020 | 7570 | 208957 | 8/19/2020 | 366106 |
|  |  | Scenario 4 | 4/24/2020 | 2721 | 39658 | 7/14/2020 | 85629 |
| Indiana | 14399 | Scenario 1 | 4/25/2020 | 718 | 14399 | 7/8/2020 | 31073 |
|  |  | Scenario 2 | 5/5/2020 | 879 | 23485 | 7/17/2020 | 42433 |
|  |  | Scenario 3 | 5/18/2020 | 1772 | 47848 | 8/4/2020 | 85601 |
|  |  | Scenario 4 | 4/25/2020 | 718 | 14399 | 6/28/2020 | 23592 |
| Iowa | 5092 | Scenario 1 | 5/23/2020 | 1129 | 30572 | 8/5/2020 | 54509 |
|  |  | Scenario 2 | 5/27/2020 | 1789 | 48976.5 | 8/13/2020 | 86478 |
|  |  | Scenario 3 | 6/1/2020 | 3015 | 83615 | 8/21/2020 | 145783 |
|  |  | Scenario 4 | 4/25/2020 | 647 | 5092 | 7/15/2020 | 21752 |
| Kansas | 3135 | Scenario 1 | 5/18/2020 | 425 | 11730.5 | 7/23/2020 | 20472 |
|  |  | Scenario 2 | 5/23/2020 | 639 | 17533 | 7/31/2020 | 30799 |
|  |  | Scenario 3 | 5/28/2020 | 1139 | 31310.5 | 8/10/2020 | 54984 |
|  |  | Scenario 4 | 4/23/2020 | 390 | 2721 | 7/8/2020 | 10699 |
| Kentucky | 3915 | Scenario 1 | 4/10/2020 | 352 | 1693 | 7/3/2020 | 9861 |
|  |  | Scenario 2 | 4/10/2020 | 352 | 1693 | 7/12/2020 | 13579 |
|  |  | Scenario 3 | 5/20/2020 | 553 | 15283.5 | 7/27/2020 | 26665 |
|  |  | Scenario 4 | 4/10/2020 | 352 | 1693 | 6/23/2020 | 7311 |
| Louisiana | 26512 | Scenario 1 | 4/2/2020 | 2725 | 9149 | 6/16/2020 | 30401 |
|  |  | Scenario 2 | 4/2/2020 | 2725 | 9149 | 6/20/2020 | 32042 |
|  |  | Scenario 3 | 4/2/2020 | 2725 | 9149 | 6/26/2020 | 35146 |
|  |  | Scenario 4 | 4/2/2020 | 2725 | 9149 | 6/12/2020 | 29268 |
| Maine | 965 | Scenario 1 | 4/2/2020 | 73 | 376 | 5/27/2020 | 1201 |
|  |  | Scenario 2 | 4/2/2020 | 73 | 376 | 6/1/2020 | 1309 |
|  |  | Scenario 3 | 4/2/2020 | 73 | 376 | 6/6/2020 | 1461 |
|  |  | Scenario 4 | 4/2/2020 | 73 | 376 | 5/21/2020 | 1101 |
| Maryland | 17766 | Scenario 1 | 4/8/2020 | 1158 | 5529 | 7/20/2020 | 54423 |
|  |  | Scenario 2 | 5/15/2020 | 1857 | 50749.5 | 8/1/2020 | 89784 |
|  |  | Scenario 3 | 5/24/2020 | 3631 | 99434 | 8/14/2020 | 175489 |
|  |  | Scenario 4 | 4/8/2020 | 1158 | 5529 | 7/7/2020 | 36188 |
| Massachusetts | 53348 | Scenario 1 | 4/24/2020 | 4946 | 50969 | 8/6/2020 | 238370 |
|  |  | Scenario 2 | 5/20/2020 | 7714 | 214408 | 8/16/2020 | 373152 |
|  |  | Scenario 3 | 5/26/2020 | 13573 | 374504 | 8/26/2020 | 656458 |
|  |  | Scenario 4 | 4/24/2020 | 4946 | 50969 | 7/22/2020 | 136457 |
| Michigan | 37074 | Scenario 1 | 4/3/2020 | 1953 | 12744 | 6/30/2020 | 51614 |
|  |  | Scenario 2 | 4/3/2020 | 1953 | 12744 | 7/4/2020 | 57180 |
|  |  | Scenario 3 | 4/3/2020 | 1953 | 12744 | 7/13/2020 | 72172 |
|  |  | Scenario 4 | 4/3/2020 | 1953 | 12744 | 6/24/2020 | 46836 |
| Minnesota | 3446 | Scenario 1 | 5/14/2020 | 353 | 9585.5 | 7/18/2020 | 16991 |
|  |  | Scenario 2 | 5/20/2020 | 571 | 15631 | 7/28/2020 | 27527 |
|  |  | Scenario 3 | 5/27/2020 | 1042 | 28390 | 8/8/2020 | 50311 |
|  |  | Scenario 4 | 4/17/2020 | 261 | 2758 | 7/3/2020 | 9361 |
| Mississippi | 5718 | Scenario 1 | 4/19/2020 | 300 | 4274 | 7/6/2020 | 14132 |
|  |  | Scenario 2 | 5/10/2020 | 426 | 11534 | 7/15/2020 | 20497 |
|  |  | Scenario 3 | 5/21/2020 | 859 | 23626 | 7/31/2020 | 41498 |
|  |  | Scenario 4 | 4/19/2020 | 300 | 4274 | 6/25/2020 | 10367 |
| Missouri | 6935 | Scenario 1 | 4/10/2020 | 465 | 3897 | 6/20/2020 | 10274 |
|  |  | Scenario 2 | 4/10/2020 | 465 | 3897 | 6/25/2020 | 11660 |
|  |  | Scenario 3 | 4/10/2020 | 465 | 3897 | 7/5/2020 | 15727 |
|  |  | Scenario 4 | 4/10/2020 | 465 | 3897 | 6/13/2020 | 9083 |
| Montana | 445 | Scenario 1 | 4/2/2020 | 33 | 241 | 5/4/2020 | 455 |
|  |  | Scenario 2 | 4/2/2020 | 33 | 241 | 5/5/2020 | 457 |
|  |  | Scenario 3 | 4/2/2020 | 33 | 241 | 5/9/2020 | 466 |
|  |  | Scenario 4 | 4/2/2020 | 33 | 241 | 5/2/2020 | 452 |
| Nebraska | 2719 | Scenario 1 | 5/24/2020 | 631 | 17404 | 8/1/2020 | 30460 |
|  |  | Scenario 2 | 5/27/2020 | 947 | 26079.5 | 8/7/2020 | 45673 |
|  |  | Scenario 3 | 6/1/2020 | 1694 | 46693 | 8/16/2020 | 81856 |
|  |  | Scenario 4 | 4/23/2020 | 389 | 2202 | 7/13/2020 | 12837 |
| Nevada | 4539 | Scenario 1 | 4/17/2020 | 310 | 3524 | 6/19/2020 | 7179 |
|  |  | Scenario 2 | 4/17/2020 | 310 | 3524 | 6/26/2020 | 8545 |
|  |  | Scenario 3 | 4/17/2020 | 310 | 3524 | 7/9/2020 | 13025 |
|  |  | Scenario 4 | 4/17/2020 | 310 | 3524 | 6/11/2020 | 6087 |
| New Hampshire | 1797 | Scenario 1 | 4/15/2020 | 217 | 1139 | 6/19/2020 | 3524 |
|  |  | Scenario 2 | 4/15/2020 | 217 | 1139 | 6/28/2020 | 4660 |
|  |  | Scenario 3 | 4/15/2020 | 217 | 1139 | 7/16/2020 | 9310 |
|  |  | Scenario 4 | 4/15/2020 | 217 | 1139 | 6/10/2020 | 2752 |
| New Jersey | 105498 | Scenario 1 | 4/3/2020 | 4305 | 29895 | 7/11/2020 | 157502 |
|  |  | Scenario 2 | 4/3/2020 | 4305 | 29895 | 7/17/2020 | 182401 |
|  |  | Scenario 3 | 5/5/2020 | 5673 | 155549 | 7/29/2020 | 270798 |
|  |  | Scenario 4 | 4/3/2020 | 4305 | 29895 | 7/4/2020 | 139015 |
| New Mexico | 2660 | Scenario 1 | 4/22/2020 | 239 | 2210 | 7/11/2020 | 10120 |
|  |  | Scenario 2 | 5/15/2020 | 288 | 7917 | 7/18/2020 | 13833 |
|  |  | Scenario 3 | 5/24/2020 | 556 | 15385 | 7/31/2020 | 26815 |
|  |  | Scenario 4 | 4/22/2020 | 239 | 2210 | 6/27/2020 | 6260 |
| New York | 282143 | Scenario 1 | 4/15/2020 | 11434 | 214454 | 7/17/2020 | 407041 |
|  |  | Scenario 2 | 4/15/2020 | 11434 | 214454 | 7/22/2020 | 460684 |
|  |  | Scenario 3 | 5/1/2020 | 12783 | 350634 | 8/1/2020 | 616569 |
|  |  | Scenario 4 | 4/15/2020 | 11434 | 214454 | 7/9/2020 | 362008 |
| North Carolina | 8768 | Scenario 1 | 4/25/2020 | 478 | 8768 | 7/10/2020 | 22742 |
|  |  | Scenario 2 | 5/11/2020 | 685 | 18753 | 7/20/2020 | 33060 |
|  |  | Scenario 3 | 5/21/2020 | 1404 | 38494 | 8/5/2020 | 67880 |
|  |  | Scenario 4 | 4/25/2020 | 478 | 8768 | 6/28/2020 | 15825 |
| North Dakota | 803 | Scenario 1 | 4/18/2020 | 135 | 528 | 6/27/2020 | 2553 |
|  |  | Scenario 2 | 4/18/2020 | 135 | 528 | 7/8/2020 | 4049 |
|  |  | Scenario 3 | 5/22/2020 | 152 | 4117 | 7/20/2020 | 7262 |
|  |  | Scenario 4 | 4/18/2020 | 135 | 528 | 6/14/2020 | 1712 |
| Northern Mariana Islands | 14 | Scenario 1 | 4/1/2020 | 4 | 6 | 4/18/2020 | 14 |
|  |  | Scenario 2 | 4/1/2020 | 4 | 6 | 4/18/2020 | 14 |
|  |  | Scenario 3 | 4/1/2020 | 4 | 6 | 4/18/2020 | 14 |
|  |  | Scenario 4 | 4/1/2020 | 4 | 6 | 4/18/2020 | 14 |
| Ohio | 15587 | Scenario 1 | 4/19/2020 | 1380 | 11602 | 7/3/2020 | 28284 |
|  |  | Scenario 2 | 4/19/2020 | 1380 | 11602 | 7/10/2020 | 34756 |
|  |  | Scenario 3 | 4/19/2020 | 1380 | 11602 | 7/28/2020 | 65653 |
|  |  | Scenario 4 | 4/19/2020 | 1380 | 11602 | 6/28/2020 | 24401 |
| Oklahoma | 3194 | Scenario 1 | 4/4/2020 | 171 | 1161 | 6/13/2020 | 4699 |
|  |  | Scenario 2 | 4/4/2020 | 171 | 1161 | 6/19/2020 | 5460 |
|  |  | Scenario 3 | 4/4/2020 | 171 | 1161 | 7/1/2020 | 7790 |
|  |  | Scenario 4 | 4/4/2020 | 171 | 1161 | 6/7/2020 | 4185 |
| Oregon | 2253 | Scenario 1 | 4/5/2020 | 169 | 1068 | 6/8/2020 | 3113 |
|  |  | Scenario 2 | 4/5/2020 | 169 | 1068 | 6/13/2020 | 3543 |
|  |  | Scenario 3 | 4/5/2020 | 169 | 1068 | 6/23/2020 | 4611 |
|  |  | Scenario 4 | 4/5/2020 | 169 | 1068 | 6/1/2020 | 2797 |
| Pennsylvania | 41153 | Scenario 1 | 4/23/2020 | 2297 | 38379 | 7/9/2020 | 71072 |
|  |  | Scenario 2 | 4/23/2020 | 2297 | 38379 | 7/16/2020 | 86734 |
|  |  | Scenario 3 | 5/9/2020 | 3091 | 77944 | 8/1/2020 | 149182 |
|  |  | Scenario 4 | 4/23/2020 | 2297 | 38379 | 7/4/2020 | 62050 |
| Puerto Rico | 1307 | Scenario 1 | 4/4/2020 | 136 | 452 | 5/29/2020 | 1602 |
|  |  | Scenario 2 | 4/4/2020 | 136 | 452 | 6/2/2020 | 1716 |
|  |  | Scenario 3 | 4/4/2020 | 136 | 452 | 6/7/2020 | 1895 |
|  |  | Scenario 4 | 4/4/2020 | 136 | 452 | 5/24/2020 | 1503 |
| Rhode Island | 7129 | Scenario 1 | 4/17/2020 | 648 | 4177 | 7/18/2020 | 27428 |
|  |  | Scenario 2 | 5/18/2020 | 942 | 25752 | 7/30/2020 | 45537 |
|  |  | Scenario 3 | 5/25/2020 | 1726 | 47341 | 8/10/2020 | 83405 |
|  |  | Scenario 4 | 4/17/2020 | 648 | 4177 | 7/5/2020 | 17204 |
| South Carolina | 5253 | Scenario 1 | 4/9/2020 | 376 | 2793 | 6/21/2020 | 8451 |
|  |  | Scenario 2 | 4/9/2020 | 376 | 2793 | 6/28/2020 | 10170 |
|  |  | Scenario 3 | 4/9/2020 | 376 | 2793 | 7/9/2020 | 14808 |
|  |  | Scenario_4 | 4/9/2020 | 376 | 2793 | 6/12/2020 | 7138 |
| South Dakota | 2147 | Scenario 1 | 4/15/2020 | 180 | 1168 | 6/26/2020 | 4932 |
|  |  | Scenario 2 | 4/15/2020 | 180 | 1168 | 7/5/2020 | 6863 |
|  |  | Scenario 3 | 5/20/2020 | 304 | 8387 | 7/23/2020 | 14616 |
|  |  | Scenario 4 | 4/15/2020 | 180 | 1168 | 6/16/2020 | 3718 |
| Tennessee | 9189 | Scenario 1 | 4/23/2020 | 872 | 8266 | 7/21/2020 | 36731 |
|  |  | Scenario 2 | 5/18/2020 | 1312 | 35972 | 8/1/2020 | 63334 |
|  |  | Scenario 3 | 5/27/2020 | 2931 | 79412 | 8/16/2020 | 141634 |
|  |  | Scenario 4 | 4/23/2020 | 872 | 8266 | 7/2/2020 | 18798 |
| Texas | 24153 | Scenario 1 | 4/9/2020 | 1431 | 11208 | 7/4/2020 | 41289 |
|  |  | Scenario 2 | 4/9/2020 | 1431 | 11208 | 7/11/2020 | 49368 |
|  |  | Scenario 3 | 5/9/2020 | 1736 | 45163 | 7/26/2020 | 83597 |
|  |  | Scenario 4 | 4/9/2020 | 1431 | 11208 | 6/29/2020 | 35839 |
| Utah | 3948 | Scenario 1 | 4/10/2020 | 247 | 2103 | 6/25/2020 | 7683 |
|  |  | Scenario 2 | 4/10/2020 | 247 | 2103 | 7/3/2020 | 9874 |
|  |  | Scenario 3 | 5/16/2020 | 410 | 11201 | 7/21/2020 | 19749 |
|  |  | Scenario 4 | 4/10/2020 | 247 | 2103 | 6/16/2020 | 6135 |
| Vermont | 843 | Scenario 1 | 4/4/2020 | 72 | 461 | 5/14/2020 | 895 |
|  |  | Scenario 2 | 4/4/2020 | 72 | 461 | 5/16/2020 | 904 |
|  |  | Scenario 3 | 4/4/2020 | 72 | 461 | 5/20/2020 | 945 |
|  |  | Scenario 4 | 4/4/2020 | 72 | 461 | 5/12/2020 | 880 |
| Virginia | 12366 | Scenario 1 | 5/9/2020 | 895 | 24037 | 7/20/2020 | 43181 |
|  |  | Scenario 2 | 5/17/2020 | 1454 | 39570 | 8/1/2020 | 70237 |
|  |  | Scenario 3 | 5/25/2020 | 2660 | 74198 | 8/13/2020 | 128592 |
|  |  | Scenario 4 | 4/25/2020 | 772 | 12366 | 7/7/2020 | 27688 |
| Virgin Islands | 55 | Scenario 1 | 3/24/2020 | 10 | 17 | 4/25/2020 | 55 |
|  |  | Scenario 2 | 3/24/2020 | 10 | 17 | 4/25/2020 | 55 |
|  |  | Scenario 3 | 3/24/2020 | 10 | 17 | 4/25/2020 | 55 |
|  |  | Scenario 4 | 3/24/2020 | 10 | 17 | 4/25/2020 | 55 |
| Washington | 13319 | Scenario 1 | 4/2/2020 | 781 | 6389 | 6/15/2020 | 16286 |
|  |  | Scenario 2 | 4/2/2020 | 781 | 6389 | 6/19/2020 | 17450 |
|  |  | Scenario 3 | 4/2/2020 | 781 | 6389 | 6/26/2020 | 19910 |
|  |  | Scenario 4 | 4/2/2020 | 781 | 6389 | 6/9/2020 | 15260 |
| West Virginia | 1010 | Scenario 1 | 4/26/2020 | 124 | 1134 | 6/13/2020 | 2016 |
|  |  | Scenario 2 | 4/26/2020 | 155 | 1165 | 6/24/2020 | 2930 |
|  |  | Scenario 3 | 4/26/2020 | 202 | 1212 | 7/16/2020 | 7242 |
|  |  | Scenario 4 | 4/19/2020 | 105 | 890 | 6/2/2020 | 1489 |
| Wisconsin | 5687 | Scenario 1 | 4/25/2020 | 331 | 5687 | 7/5/2020 | 13919 |
|  |  | Scenario 2 | 5/9/2020 | 412 | 11238 | 7/15/2020 | 19836 |
|  |  | Scenario 3 | 5/20/2020 | 847 | 23199.5 | 7/31/2020 | 40891 |
|  |  | Scenario 4 | 4/25/2020 | 331 | 5687 | 6/22/2020 | 9649 |
| Wyoming | 491 | Scenario 1 | 4/21/2020 | 126 | 443 | 6/13/2020 | 1127 |
|  |  | Scenario 2 | 4/21/2020 | 126 | 443 | 6/21/2020 | 1477 |
|  |  | Scenario 3 | 4/21/2020 | 126 | 443 | 7/4/2020 | 2450 |
|  |  | Scenario 4 | 4/21/2020 | 126 | 443 | 6/6/2020 | 884 |

**Table S4.** Estimated time varying intervention measures of US and top 10 states with the largest number of end cases.

| Date | US | California | Connecticut | Illinois | Iowa | Maryland | Massachusetts | Michigan | New Jersey | New York | Pennsylvania |
| --- | --- | --- | --- | --- | --- | --- | --- | --- | --- | --- | --- |
| 4/15/2020 | 0.5407 | 0.5463 | 0.5152 | 0.5190 | 0.5044 | 0.5190 | 0.5169 | 0.5572 | 0.5282 | 0.5748 | 0.5258 |
| 4/16/2020 | 0.5513 | 0.5461 | 0.5252 | 0.5238 | 0.5111 | 0.5189 | 0.5222 | 0.5670 | 0.5457 | 0.5674 | 0.5350 |
| 4/17/2020 | 0.5561 | 0.5600 | 0.5167 | 0.5329 | 0.5113 | 0.5182 | 0.5191 | 0.5970 | 0.5459 | 0.5705 | 0.5379 |
| 4/18/2020 | 0.5591 | 0.5592 | 0.5187 | 0.5293 | 0.5060 | 0.5144 | 0.5182 | 0.6026 | 0.5555 | 0.5916 | 0.5361 |
| 4/19/2020 | 0.5625 | 0.5609 | 0.5307 | 0.5290 | 0.5049 | 0.5144 | 0.5212 | 0.6236 | 0.5708 | 0.6162 | 0.5336 |
| 4/20/2020 | 0.5763 | 0.5827 | 0.5631 | 0.5394 | 0.4899 | 0.5265 | 0.5271 | 0.6495 | 0.5732 | 0.6392 | 0.5480 |
| 4/21/2020 | 0.5961 | 0.5581 | 0.5271 | 0.5586 | 0.4889 | 0.5264 | 0.5934 | 0.6749 | 0.5791 | 0.6600 | 0.5738 |
| 4/22/2020 | 0.6096 | 0.5496 | 0.5407 | 0.5627 | 0.4747 | 0.5400 | 0.5601 | 0.6881 | 0.5854 | 0.6769 | 0.5889 |
| 4/23/2020 | 0.6173 | 0.5476 | 0.5200 | 0.5506 | 0.4980 | 0.5579 | 0.5599 | 0.6702 | 0.5932 | 0.6918 | 0.6183 |
| 4/24/2020 | 0.6240 | 0.5417 | 0.5308 | 0.5483 | 0.5133 | 0.5485 | 0.5424 | 0.6516 | 0.5943 | 0.6995 | 0.5871 |
| 4/25/2020 | 0.6183 | 0.5470 | 0.5540 | 0.5313 | 0.5058 | 0.5421 | 0.5122 | 0.6390 | 0.6238 | 0.6908 | 0.5765 |
| 4/26/2020 | 0.6183 | 0.5781 | 0.5872 | 0.5313 | 0.5058 | 0.5421 | 0.5131 | 0.6696 | 0.6348 | 0.6908 | 0.6024 |
| 4/27/2020 | 0.6237 | 0.5947 | 0.5908 | 0.5322 | 0.5058 | 0.5421 | 0.5167 | 0.6771 | 0.6451 | 0.6908 | 0.6182 |
| 4/28/2020 | 0.6311 | 0.6041 | 0.5926 | 0.5362 | 0.5058 | 0.5421 | 0.5195 | 0.6797 | 0.6536 | 0.6908 | 0.6283 |
| 4/29/2020 | 0.6385 | 0.6112 | 0.5998 | 0.5410 | 0.5058 | 0.5421 | 0.5195 | 0.6834 | 0.6624 | 0.6908 | 0.6293 |
| 4/30/2020 | 0.6456 | 0.6201 | 0.6064 | 0.5454 | 0.5058 | 0.5421 | 0.5235 | 0.6921 | 0.6713 | 0.6908 | 0.6346 |
| 5/1/2020 | 0.6535 | 0.6287 | 0.6142 | 0.5501 | 0.5058 | 0.5477 | 0.5246 | 0.6992 | 0.6789 | 0.6908 | 0.6436 |
| 5/2/2020 | 0.6617 | 0.6376 | 0.6199 | 0.5546 | 0.5058 | 0.5538 | 0.5246 | 0.7053 | 0.6861 | 0.6938 | 0.6524 |
| 5/3/2020 | 0.6692 | 0.6457 | 0.6262 | 0.5595 | 0.5058 | 0.5594 | 0.5288 | 0.7112 | 0.6929 | 0.7010 | 0.6602 |
| 5/4/2020 | 0.6767 | 0.6539 | 0.6339 | 0.5655 | 0.5058 | 0.5658 | 0.5324 | 0.7192 | 0.6995 | 0.7097 | 0.6672 |
| 5/5/2020 | 0.6842 | 0.6617 | 0.6413 | 0.5721 | 0.5073 | 0.5724 | 0.5354 | 0.7265 | 0.7067 | 0.7174 | 0.6748 |
| 5/6/2020 | 0.6914 | 0.6693 | 0.6492 | 0.5787 | 0.5093 | 0.5790 | 0.5412 | 0.7336 | 0.7145 | 0.7250 | 0.6823 |
| 5/7/2020 | 0.6984 | 0.6769 | 0.6568 | 0.5864 | 0.5116 | 0.5866 | 0.5471 | 0.7402 | 0.7221 | 0.7324 | 0.6895 |
| 5/8/2020 | 0.7055 | 0.6842 | 0.6645 | 0.5950 | 0.5133 | 0.5952 | 0.5529 | 0.7485 | 0.7292 | 0.7389 | 0.6964 |
| 5/9/2020 | 0.7135 | 0.6913 | 0.6722 | 0.6035 | 0.5150 | 0.6038 | 0.5584 | 0.7617 | 0.7360 | 0.7463 | 0.7030 |
| 5/10/2020 | 0.7211 | 0.6983 | 0.6798 | 0.6120 | 0.5171 | 0.6123 | 0.5646 | 0.7782 | 0.7424 | 0.7580 | 0.7110 |
| 5/11/2020 | 0.7284 | 0.7055 | 0.6871 | 0.6204 | 0.5212 | 0.6207 | 0.5713 | 0.7971 | 0.7513 | 0.7748 | 0.7188 |
| 5/12/2020 | 0.7352 | 0.7134 | 0.6942 | 0.6288 | 0.5248 | 0.6291 | 0.5778 | 0.8170 | 0.7664 | 0.7925 | 0.7261 |
| 5/13/2020 | 0.7417 | 0.7211 | 0.7011 | 0.6370 | 0.5283 | 0.6374 | 0.5852 | 0.8373 | 0.7829 | 0.8119 | 0.7331 |
| 5/14/2020 | 0.7503 | 0.7283 | 0.7088 | 0.6451 | 0.5316 | 0.6454 | 0.5936 | 0.8558 | 0.8015 | 0.8329 | 0.7397 |
| 5/15/2020 | 0.7645 | 0.7352 | 0.7166 | 0.6531 | 0.5351 | 0.6534 | 0.6021 | 0.8722 | 0.8219 | 0.8519 | 0.7475 |
| 5/16/2020 | 0.7811 | 0.7416 | 0.7241 | 0.6610 | 0.5412 | 0.6613 | 0.6105 | 0.8866 | 0.8417 | 0.8688 | 0.7592 |
| 5/17/2020 | 0.7994 | 0.7503 | 0.7312 | 0.6687 | 0.5472 | 0.6691 | 0.6189 | 0.8993 | 0.8597 | 0.8835 | 0.7761 |
| 5/18/2020 | 0.8196 | 0.7645 | 0.7379 | 0.6763 | 0.5531 | 0.6766 | 0.6273 | 0.9095 | 0.8756 | 0.8967 | 0.7937 |
| 5/19/2020 | 0.8397 | 0.7811 | 0.7450 | 0.6837 | 0.5587 | 0.6841 | 0.6356 | 0.9175 | 0.8898 | 0.9076 | 0.8131 |
| 5/20/2020 | 0.8579 | 0.7994 | 0.7549 | 0.6909 | 0.5650 | 0.6912 | 0.6437 | 0.9244 | 0.9020 | 0.9158 | 0.8339 |
| 5/21/2020 | 0.8740 | 0.8195 | 0.7716 | 0.6979 | 0.5716 | 0.6982 | 0.6517 | 0.9313 | 0.9114 | 0.9230 | 0.8526 |
| 5/22/2020 | 0.8884 | 0.8397 | 0.7884 | 0.7050 | 0.5781 | 0.7053 | 0.6596 | 0.9378 | 0.9192 | 0.9299 | 0.8694 |
| 5/23/2020 | 0.9008 | 0.8578 | 0.8072 | 0.7130 | 0.5855 | 0.7133 | 0.6674 | 0.9433 | 0.9259 | 0.9364 | 0.8843 |
| 5/24/2020 | 0.9105 | 0.8740 | 0.8282 | 0.7206 | 0.5939 | 0.7209 | 0.6750 | 0.9479 | 0.9328 | 0.9421 | 0.8973 |
| 5/25/2020 | 0.9184 | 0.8883 | 0.8474 | 0.7279 | 0.6025 | 0.7282 | 0.6824 | 0.9517 | 0.9391 | 0.9469 | 0.9080 |
| 5/26/2020 | 0.9251 | 0.9007 | 0.8647 | 0.7348 | 0.6109 | 0.7351 | 0.6897 | 0.9548 | 0.9443 | 0.9508 | 0.9162 |
| 5/27/2020 | 0.9321 | 0.9105 | 0.8800 | 0.7413 | 0.6193 | 0.7415 | 0.6967 | 0.9574 | 0.9487 | 0.9541 | 0.9233 |
| 5/28/2020 | 0.9384 | 0.9184 | 0.8935 | 0.7497 | 0.6277 | 0.7501 | 0.7036 | 0.9595 | 0.9524 | 0.9568 | 0.9302 |
| 5/29/2020 | 0.9438 | 0.9251 | 0.9050 | 0.7634 | 0.6360 | 0.7643 | 0.7116 | 0.9613 | 0.9554 | 0.9590 | 0.9367 |
| 5/30/2020 | 0.9483 | 0.9321 | 0.9138 | 0.7801 | 0.6441 | 0.7809 | 0.7193 | 0.9627 | 0.9578 | 0.9608 | 0.9424 |
| 5/31/2020 | 0.9520 | 0.9385 | 0.9212 | 0.7983 | 0.6521 | 0.7993 | 0.7266 | 0.9639 | 0.9599 | 0.9624 | 0.9471 |

**Table S5.** Correlation coefficients between the number of cumulative cases and intervention measures in total US and individual states.

| US and State | Correlation Coefficients | State | Correlation Coefficients |
| --- | --- | --- | --- |
| US | -0.4819 | Montana | -0.3078 |
| Alabama | -0.3069 | Nebraska | -0.2822 |
| Alaska | -0.3065 | Nevada | -0.3061 |
| Arizona | -0.4008 | New Hampshire | -0.2781 |
| Arkansas | -0.2844 | New Jersey | -0.3346 |
| California | -0.4473 | New Mexico | -0.2900 |
| Colorado | -0.3011 | New York | -0.3442 |
| Connecticut | -0.2915 | North Carolina | -0.3226 |
| Delaware | -0.3007 | North Dakota | -0.2834 |
| District of Columbia | -0.3109 | Northern Mariana Islands | -0.1480 |
| Florida | -0.3243 | Ohio | -0.3056 |
| Georgia | -0.3148 | Oklahoma | -0.3179 |
| Guam | -0.2607 | Oregon | -0.3126 |
| Hawaii | -0.2679 | Pennsylvania | -0.3235 |
| Idaho | -0.2558 | Puerto Rico | -0.2613 |
| Illinois | -0.3759 | Rhode Island | -0.3018 |
| Indiana | -0.3231 | South Carolina | -0.3039 |
| Iowa | -0.2790 | South Dakota | -0.3050 |
| Kansas | -0.2886 | Tennessee | -0.2949 |
| Kentucky | -0.2975 | Texas | -0.3275 |
| Louisiana | -0.2804 | Utah | -0.3138 |
| Maine | -0.3030 | Vermont | -0.3050 |
| Maryland | -0.3115 | Virginia | -0.3091 |
| Massachusetts | -0.3521 | Virgin Islands | -0.2058 |
| Michigan | -0.3160 | Washington | -0.4197 |
| Minnesota | -0.3001 | West Virginia | -0.2490 |
| Mississippi | -0.3135 | Wisconsin | -0.3099 |
| Missouri | -0.2970 | Wyoming | -0.2232 |

## Notes

### Competing Interest Statement

The authors have declared no competing interest.

